# Ascertainment rate of novel coronavirus disease (COVID-19) in Japan

**DOI:** 10.1101/2020.03.09.20033183

**Authors:** Ryosuke Omori, Kenji Mizumoto, Hiroshi Nishiura

## Abstract

**Objective:** To estimate the ascertainment rate of novel coronavirus (COVID-19).

**Methods:** We analyzed the epidemiological dataset of confirmed cases with COVID-19 in Japan as of 28 February 2020. A statistical model was constructed to describe the heterogeneity of reporting rate by age and severity. We estimated the number of severe and non-severe cases, accounting for under-ascertainment.

**Results:** The ascertainment rate of non-severe cases was estimated at 0.44 (95% confidence interval: 0.37, 0.50), indicating that unbiased number of non-cases would be more than twice the reported count.

**Conclusions:** Severe cases are twice more likely diagnosed and reported than other cases. Considering that reported cases are usually dominated by non-severe cases, the adjusted total number of cases is also about a double of observed count. Our finding is critical in interpreting the reported data, and it is advised to interpret mild case data of COVID-19 as always under-ascertained.

**Highlights:**

- Epidemiological dataset of COVID-19 in Japan was analyzed.
- The ascertainment rate of non-severe cases was estimated at 0.44 (95% confidence interval: 0.37, 0.50).
- Severe cases are twice more likely diagnosed and reported than other cases.
- Mild cases of COVID-19 are under-ascertained.

## Introduction

As of 1 March 2020, a total of 58 countries reported at least one confirmed case of novel coronavirus disease (COVID-19), and the cumulative number of deaths reached 2977 persons across the world (WHO, 2020). To attain appropriate countermeasures, it is vital to understand current epidemiological situations of the COVID-19 epidemic.

The majority of COVID-19 cases exhibit limited severity; 81% of reported cases in China has been mild and only 16% are severe (Guan et al., 2020). It is natural that the ascertainment rate would be different between severe and non-severe cases. The present study aims to estimate the ascertainment rate of non-severe cases, employing a statistical model.

## Methods

We analyzed the epidemiological dataset of confirmed cases with COVID-19 in Japan as of 28 February 2020. The confirmatory diagnosis was made by means of reverse transcriptase polymerase chain reaction (RT-PCR). The present study specifically analyzed cases by (i) prefecture, (ii) age, and (iii) severity. Severe case was defined as (i) severe dyspnea that required oxygen support plus pneumonia or intubation or (ii) case that required management in intensive care unit.

We estimated the number of severe and non-severe cases using the ratio of non-severe to severe reported cases (Guan et al., 2020, Novel, 2020). We estimated the ascertainment rate among non-severe cases by 1/*k*, describing data generating process of both severe and non-severe generated from Poisson process with probabilities *p*_x,a_ for severe cases and *kf*_a_*p*_x,a_ for non-severe cases in age group *a* and prefecture *x*, respectively. Here *f*_a_ denotes the ratio of non-severe to severe reported case of age group *a*, as estimated from age-specific severity and incidence rate ratio in China (Guan et al., 2020, Novel, 2020). We estimate *k* and *p*_x,a_ using the loglikelihood function:

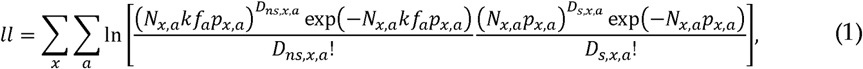

where *N*_*x,a*_, *D*_*ns,x,a*_ and *D*_*s,x,a*_ represent the population size, the observed counts of non-severe and severe cases of age group *a* in prefecture *x*, respectively.

Maximum likelihood estimates were obtained by maximizing the equation (1) and the profile likelihood-based confidence intervals were computed.

## Results

The ascertainment rate of non-severe cases, *k*, was estimated at 0.44 (95% confidence interval (CI): 0.37, 0.50). Resulting estimate of non-severe cases is shown in Figure 1A, showing along with reasonably good fit to severe case data in Figure 1B. Age-specific pattern of estimated non-severe cases was similar to that among severe cases. The largest estimated number of non-severe cases was 80 cases (95% CI: 63, 98) among those aged 50-59 years and 78 (95% CI: 61, 95) among cases aged 60-69 years, respectively. Such adjustment gives adjusted estimate of the total cases by age group.

**Figure 1.**
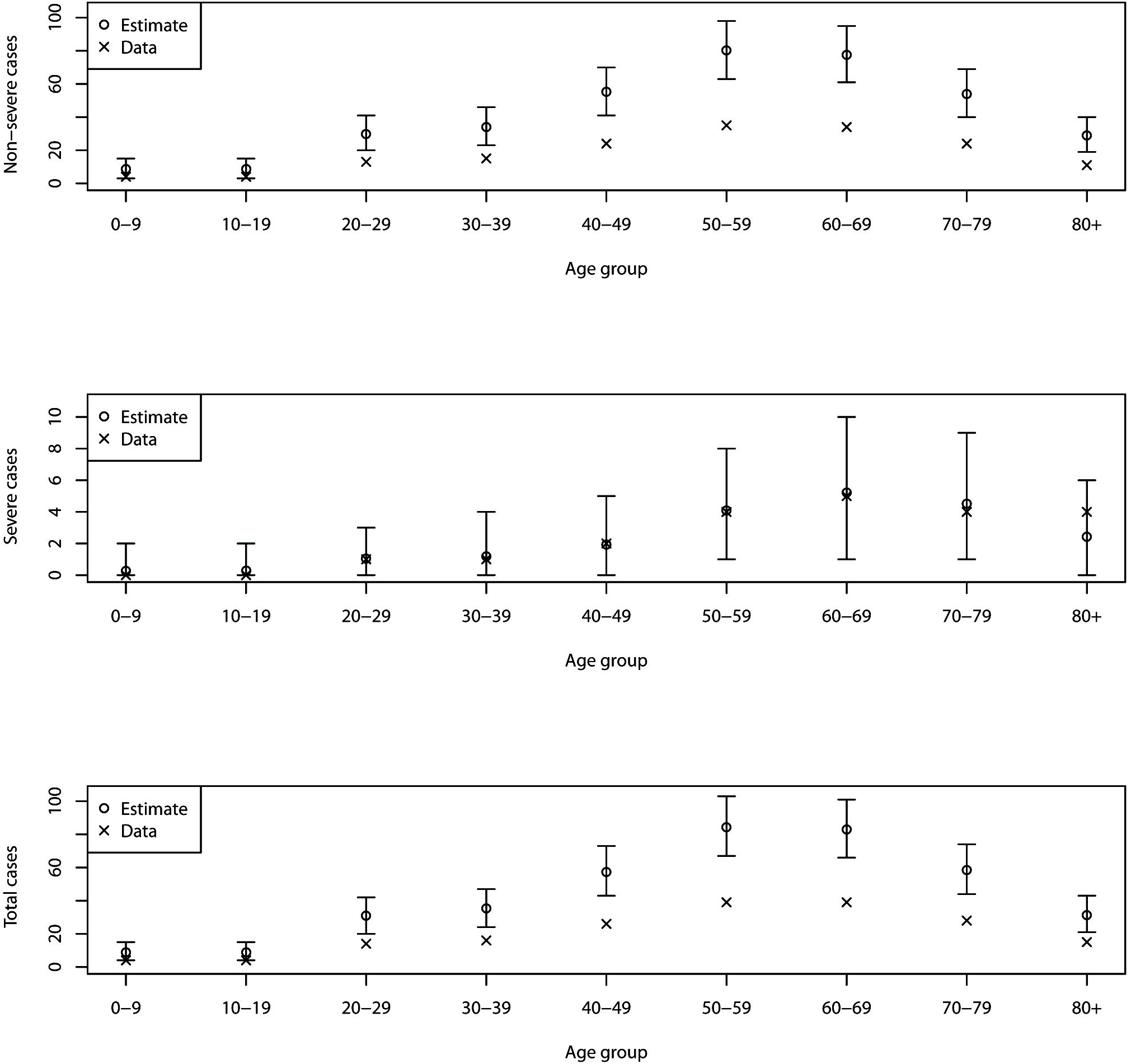
Age-specific number of novel coronavirus disease (COVID-19) cases by age group and severity. Top: non severe cases, middle: severe cases, and bottom: total cases. x-marks represent observed counts, while unfilled circles show estimated cases. Whiskers extend to lower and upper 95% confidence intervals, derived from profile likelihood.

## Discussion

The present study estimated the ascertainment-adjusted number of cases in Japan, using age-specific severe fraction of cases. We assumed that the ratio of severe to non-severe cases in a given age group is a constant and that the age-independent gap is explained by the under-diagnosis and under-reporting, estimating the ascertainment rate among non-severe cases to be 0.44.

As a take home, it must be remembered that severe cases are twice more likely diagnosed and reported than other cases. Reported cases are usually dominated by non-severe cases, and the adjusted total number of cases is about a double of observed count. Our finding is critical in interpreting the reported data, and it is advised to regard the mild case data as always under-ascertained.

In addition to the proposed adjustment, it should be noted that the ascertainment rate of severe cases needs to be additionally estimated, and such estimation requires direct measurement of the total number of cases or infected individuals by means of seroepidemiological study or other testing methods of all samples (Nishiura et al., 2020). That is, the actual total number of cases is greater than what it was adjusted in the present study. Using seroepidemiological datasets, we plan to address relevant issues in the future. Other limitations include that (i) we did not explore detailed natural history, e.g. dynamically changing symptoms over the course of infection, and underlying comorbidities, (ii) we ignored right-censored data, e.g. the time delay from illness onset to severe manifestations, for simplicity. The latter led us to underestimate the ascertainment rate. (iii) it is worth noting that the data of age dependent severity employed in our analysis is only based on the observed data in China. Considering the possibility of underreporting or biased age distribution, the nature of this age distribution may lead to underestimation.

Despite multiple future tasks, we believe that the present study successfully demonstrated that the ascertainment rate can be partly adjusted by examining age-dependent number of cases including severe cases. The proposed adjustment should be practiced in other country settings and also for other diseases.

## Data Availability

The data that support the findings of this study are available from the corresponding author upon reasonable request.

## Conflict of interest

The authors declare no conflicts of interest.

## Funding source

H.N. received funding support from Japan Agency for Medical Research and Development [grant number: JP18fk0108050] the Japan Society for the Promotion of Science (JSPS) Grants-in-Aid for Scientific Research (KAKENHI in Japanese abbreviation) grant nos. 17H04701, 17H05808, 18H04895 and 19H01074, and the Japan Science and Technology Agency (JST) Core Research for Evolutional Science and Technology (CREST) program [grant number: JPMJCR1413]. The funders had no role in study design, data collection and analysis, decision to publish, or preparation of the manuscript.

## Ethical approval

This study was based on publicly available data and did not require ethical approval.

## Notes

### Competing Interest Statement

The authors have declared no competing interest.

